# Artificial Intelligence-assisted reader evaluation in acute CT head interpretation (AI-REACT): a multireader multicase study

**DOI:** 10.1101/2025.10.17.25337471

**Authors:** Alex Novak, Ruchir Shah, Abdalá T Espinosa Morgado, Dennis Robert, Shamie Kumar, Satish Golla, Jason Oke, Kanika Bhatia, Andrea Romsauerova, Tilak Das, Irfan Ullah Akbarkhan, Laura Hunter, Nazreen Kaneez, Ana Nicolescu, Emma Kelliher, Kyle Stephenson, Shair Ali, Danielle Benson, Fiona Hunter, Ross Hunter, Benjamin Scally, Rhys Worgan, Louise Hartley, Ryan Grech, Martine Walker, Neil Mitchell, Ravi Shashikala, Alice Gibson, Hélène Matte, Shubhendu Kulshrestha, Hannah Yang, Radoslaw Rippel, Roland Amoah, Zahi Qamhawi, Thomas Millard, Avneet Gill, Michael Thompson, Josh Beck, Harsh Merchant, Ben Lockwood, Mariusz Tadeusz Grzeda, Mariapaola Narbone, Rahul Dharmadhikari, Mark Harrison, Kavitha Vimalesvaran, Jane Gooch, Nick Woznitza, Nabeeha Salik, Alan Campbell, Farhaan Khan, David J Lowe, Haris Shuaib, Sarim Ather

## Abstract

**Background:** Non-contrast CT head scans (NCCTH) are the most frequently requested cross-sectional imaging in the Emergency Department. While AI tools have been developed to detect NCCTH abnormalities, most validation studies compare AI to radiologists, with limited evidence on the impact of AI assistance for other healthcare professionals.

**Objective:** To evaluate whether an AI-powered tool improves the accuracy, speed, and confidence of general radiologists, emergency clinicians, and radiographers in detecting critical abnormalities on NCCTH, and to assess the tool’s stand-alone performance and factors influencing diagnostic accuracy and efficiency.

**Methods:** A retrospective dataset of 150 NCCTH (52 normal, 98 with critical abnormalities: intracranial haemorrhage, hypodensity, midline shift, mass effect, or skull fracture) was reviewed by 30 readers (10 radiologists, 15 emergency clinicians, 5 radiographers) from four NHS trusts. Each reader interpreted scans first unaided, then with the qER EU 2.0 AI tool, separated by a 2-week washout. Ground truth was established by consensus of two neuroradiologists. We assessed the stand-alone performance of qER and its effect on reader diagnostic accuracy, confidence, and interpretation speed.

**Results:** The qER algorithm demonstrated strong diagnostic performance across most pathology subgroups (AUC 0.821–0.976). With AI assistance, pooled reader sensitivity for critically abnormal scans increased from 82.8% to 89.7% (+6.9%, 95% CI +1.4% to +10.6%, p<0.001), and for intracranial haemorrhage from 84.6% to 91.6% (+7.0%, 95% CI +3.2% to +10.8%, p<0.001), but specificity decreased from 84.5% to 78.9% (–5.5%, 95% CI –11.0% to –0.09%, p=0.046). Reader confidence AUC did not change significantly. ED clinicians with AI achieved sensitivity comparable to unaided radiologists, with no significant change in specificity.

**Conclusion:** AI-assisted interpretation increased reader sensitivity for critical abnormalities but reduced specificity. Notably, AI assistance enabled ED clinicians to reach diagnostic sensitivity similar to unaided radiologists, supporting the potential for AI to extend the diagnostic capabilities of non-radiologists. Further prospective studies are warranted to confirm these findings in real-world settings.

**Funding:** This study was funded by Qure.ai via an NHSX Award

**Ethics:** The study has been approved by the UK Healthcare Research Authority (IRAS 310995, approved 13/12/2022). The use of anonymised retrospective NCCTH has been authorised by Oxford University Hospitals.

**Trial registration number:** <u>NCT06018545</u>.

**Research in context:** *What is already known on this topic:* AI-derived algorithms for the detection of pathological findings on non-contrast CT head (NCCTH) images have previously demonstrated strong diagnostic performance when used on retrospective datasets. AI-assisted image interpretation using these algorithms has been shown to enhance the diagnostic performance of general and neuro-radiologists *in silico*. The potential for AI to enhance the performance of less skilled readers who may encounter and be required to act on these images in clinical practice (e.g. non-specialist radiologists, emergency medicine clinicians and radiographers) is as yet untested, however.

*What this study adds:* This large multicase multireader study demonstrates that AI-assisted image interpretation may be used to enhance the *in silico* diagnostic performance of Emergency Department physicians to a level comparable to that of general radiologists.

*How this study might affect research, practice or policy:* This study raises the possibility that AI-assisted image interpretation could be used to assist non-radiologist clinicians in the safe interpretation of NCCTH scans. Further prospective research is required to test this hypothesis in clinical practice and explore the potential for AI-assisted interpretation to support safe discharge of patients with normal or low-risk scans.

## Introduction

Computerised Tomography (CT) of the head is the most common cross sectional imaging modality performed in the Emergency Department, with over 1 million scans requested annually in the UK.^1^ Increased ED attendances combined with improved CT availability and a lower clinical threshold for scanning has resulted in a significantly greater number of patients with lower acuity receiving non-contrast CT Head (NCCTH) scans, most of whom will be ultimately discharged from the ED without admission.^2^ Typically however, ED clinicians are dependent on the NCCTH report from radiologists before making a clinical decision based on their findings.^3^ This requirement places a huge demand on radiology services which are often unable to provide timely reports leading to delays in treatment, referral and discharge decisions, ultimately impacting on patient care and departmental efficiency and reducing patient flow through the Emergency Department.^4^

A number of AI-assisted image interpretation algorithms have been developed to assist clinicians in the identification of pathological findings on NCCTH scans.^5,6^ Several of these are CE-marked and FDA-approved for clinical use and are already being deployed in hospitals worldwide. Retrospective studies have indicated high accuracy in some cases, and reader studies have demonstrated the potential for AI-assisted NCCTH interpretation to improve radiologist accuracy.^7^ To date however, evidence of efficacy and impact has been focussed primarily on the performance of radiologists, and few studies have explored the potential for AI-assisted image interpretation in evaluating the impact of AI assistance with other groups of healthcare professionals who regularly review or act on NCCTH interpretations, such as ED clinicians and radiographers. ^8,9^

qER 2.0 EU is a CE (European Conformity) Class IIb AI tool for interpretation of NCCTH, which was developed using 300⍰000 retrospectively collected and labelled scans from 31 imaging centres in India and one of the largest teleradiology centres in the USA, including scans obtained from both in-hospital and outpatient radiology settings.^10^ It can detect, classify and localise intracranial haemorrhage, hypodensities suggestive of infarct, mass effect, midline shift, atrophy and skull fractures in NCCTH. If any of the target abnormalities is detected by the software, the tool provides the user with a single summary listing all the target abnormalities found by qER on the CT, followed by all slices in the scan with the overlay highlighting the location of the abnormalities (figure 1). Alternatively, if none of the target abnormalities are detected, the output will indicate that the software has analysed the image and identified no target abnormalities.^11^

**Figure 1.**
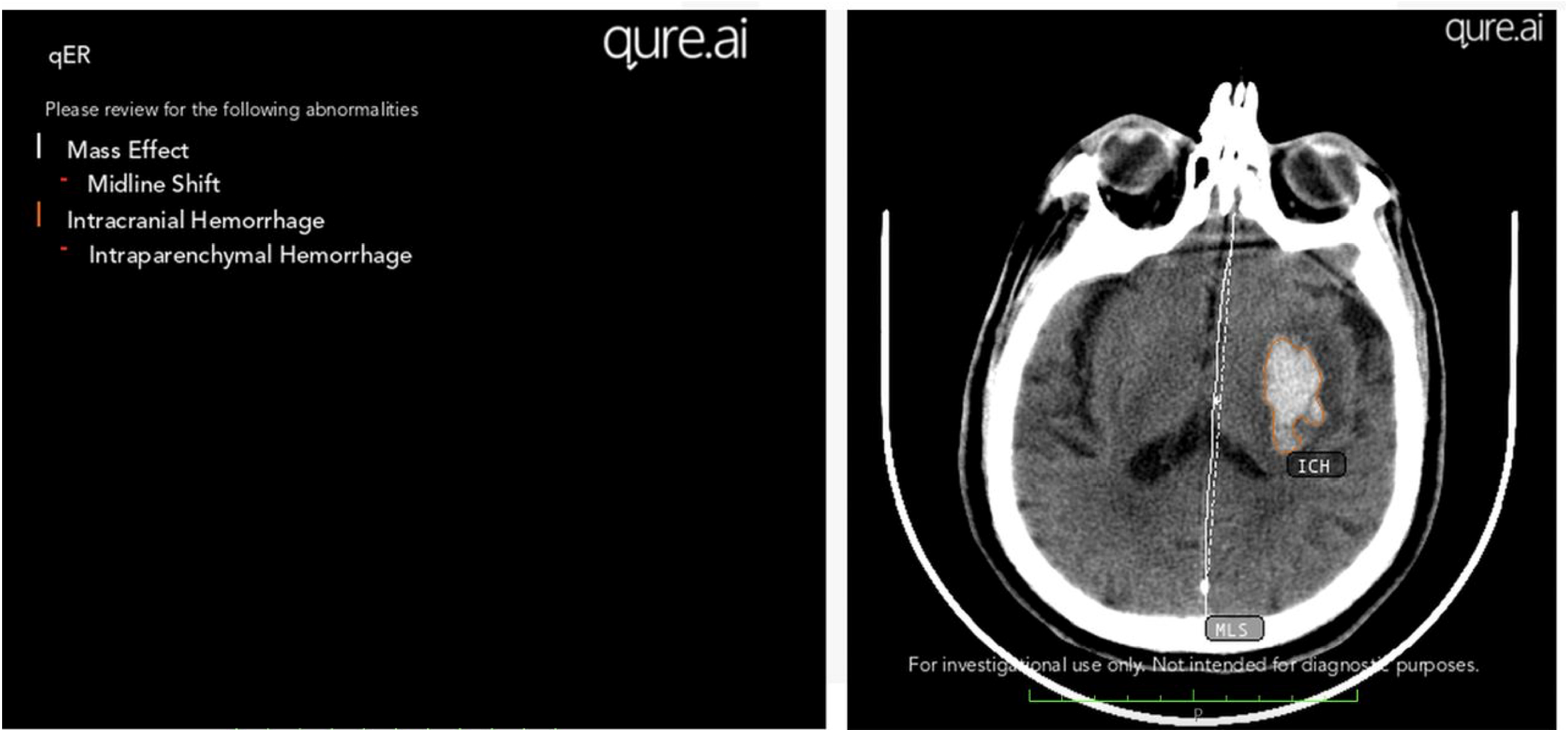
qER presents a summary of all abnormalities identified and the slice images containing those abnormalities with a localisation overlay. ICH, intracranial haemorrhage.

qER is intended to support certified radiologists and/or licensed medical practitioners for clinical decision-making. It is a support tool which, when used with original scans, may assist the clinician to improve efficiency, accuracy and turnaround time in reading NCCTH. Multiple prior studies have reported the standalone diagnostic accuracy of qER, however as yet its potential impact on the diagnostic accuracy of general radiologists, radiographers and ED clinicians has not been fully tested.^10,12-14^ Here we evaluate a deep-learning model designed to assist clinicians in the interpretation of CT head images, using clinicians and radiologists of varying seniority and experience to evaluate an image dataset which includes the full range of pathologies commonly encountered in routine emergency care.

### Study aims

1. To determine the improvement in NCCTH image interpretation accuracy of general radiologists, ED clinicians and radiographers in detecting critical abnormalities (any one or more of intracranial haemorrhage, midline shift, mass effect, skull fracture or hypodensity suggestive of infarct) with the assistance of the qER AI tool (primary).
2. To determine the stand-alone accuracy of qER for the detection of intracranial haemorrhage, hypodensity suggestive of infarct, midline shift, mass effect and skull fractures (secondary).
3. To measure the time taken by the above clinicians to evaluate scan images, and their diagnostic confidence, with and without the AI tool (secondary).
4. To explore which imaging factors influence clinicians’ reporting accuracy and efficiency, and algorithm performance, for example, category of abnormality, difficulty of image interpretation, clinician seniority and professional group (secondary).

## Methods

### Study design and participants

We undertook a fully-crossed paired multireader multicase (MRMC) study as per our previously published study protocol.^15^ 150 Non-contrast CT Head scans of ED patients aged 18 years or above were retrospectively identified by the clinical and Picture Archiving and Communication System (PACS)/ Information Technology (IT) team by searching the Radiology Information System at Oxford University Hospitals NHS Foundation Trust (see Figure S1 in supplementary material). The dataset was selected using existing clinical radiology reports to contain 60 control scans and 90 abnormal scans, including a minimum of 10 scans containing each of the following 9 defined ‘critical abnormalities’: extradural haemorrhage, subdural haemorrhage, subarachnoid haemorrhage, intraparenchymal haemorrhage, intraventricular haemorrhage, hypodensity suggestive of infarct, midline shift, mass effect, and skull fractures.

A summary of the process used to create the dataset is summarised in figure S2 in supplementary material. For the purposes of case selection, the existing clinical radiology reports were used to determine whether a given scan contains an abnormality of interest. To reduce selection bias, consecutive scans were reviewed and all scans which fitted the inclusion and exclusion criteria were included until the specific case number totals were reached. Each case was derived from a different patient. In addition to the prespecified pathology subgroups, image inclusion criteria is summarized as follows:

### Inclusion Criteria

- Individuals undergoing NCCTH in the ED
- Age ≥18 years
- Non-contrast axial CT scan series with consistently spaced axial slices
- Soft reconstruction kernel covering the complete brain
- Maximum slice thickness of 6mm.

### Exclusion Criteria

Following features which are known to cause inaccurate outputs from the qER AI:

- Scans with obvious postoperative defects, or from patients who previously underwent brain surgery.
- Scans with artefacts such as burr holes, shunts or clips.
- Scans containing metal artefacts.

## Establishing a Reference Standard

To establish a reference standard, two consultant neuroradiologists independently reviewed the CT images in the dataset and recorded the presence or absence of the 9 target abnormalities for each scan. In the case of disagreement, a third senior neuroradiologist’s opinion was sought for arbitration. A difficulty score was assigned to each pathological finding by the two ground truthers using a 5-point Likert scale, and where there was disagreement, the mean score was taken.

### Image inferencing

All images were inferenced by the qER algorithm, and the resulting secondary capture images stored as a separate dataset. The qER output was presented as an additional series with a notification to suggest the presence or absence of a target abnormality as the first image of the series and segmentation of the abnormal areas identified overlaid on the scan images.

### Reader participants

30 readers were recruited from the following four hospital Trusts: Guy’s & St Thomas NHS Foundation Trust, Northumbria Healthcare NHS Foundation Trust, NHS Greater Glasgow and Clyde and Oxford University Hospitals NHS Foundation Trust. The composition and inclusion/exclusion criteria for the readers are summarised as follows:

### Readers

#### Emergency Medicine

- 5 consultants
- 5 registrars ST3-6
- 5 juniors F1-ST2)

#### General radiologist

- 5 consultants
- 5 registrars (ST3-6)

#### Radiographers

- 5 CT Radiographers

### Inclusion Criteria

#### Clinicians who review NCCTH as part of their clinical practice including

- General Radiologists
- Radiographers
- Emergency Medicine Doctors

### Exclusion Criteria

- Neuroradiologists
- Non-radiologist readers:
- Clinicians with previous formal postgraduate CT reporting training
- Emergency medicine readers:
- Clinicians with previous career in radiology or neurosurgery to registrar level or above

### Reader phase 1 and 2

Reader recruitment was undertaken by each principal investigator for their own site via email and in person. 5 modules each containing 30 cases were created and uploaded to a secure online DICOM viewer (www.RAIQC.com). All 30 readers undertook a brief training module including 5 practice cases to familiarise themselves with the platform and study requirements, then proceeded to review all 150 cases over a 4-week period using a laptop or PC. For each scan, the readers recorded whether they identified any of the nine critical abnormalities as being present, providing a confidence rating for each of their findings on a 10-point Likert scale. The time taken for each scan interpretation was automatically recorded. The order of the cases was randomised for each reader at each phase, and readers were blinded to the number of abnormal and normal cases in the study.

In the first phase all readers reviewed all 150 scans, blinded to the ground truth and without AI assistance. Following a 2-week washout period to mitigate recall bias readers undertook the second phase, where they reviewed all scans again in a randomised order, remaining blinded to the ground truth, but this time with access to the results from the qER tool. Figure 2 includes screenshots from the RAIQC modules as presented to the readers.

**Figure 2.**
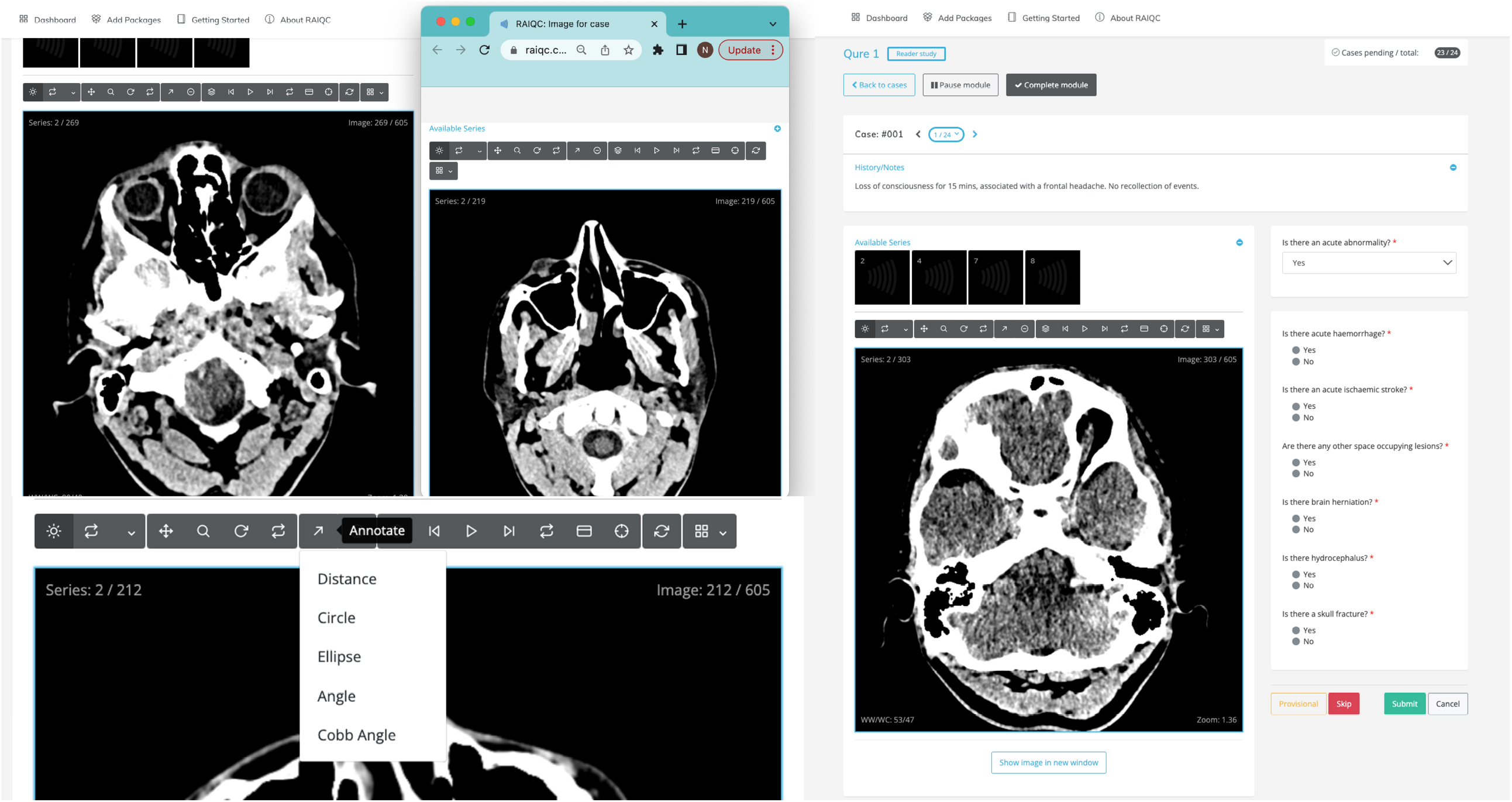
Screenshots of reader study online module interface as presented to clinical readers. (A) Readers were asked to state their confidence in each identification, on a quasi-continuous scale from 1 to 10. (B) Ground truthers were asked to rate the difficulty of detection for each pathological finding on a Likert scale from 1 to 5. (C) The artificial intelligence output was presented as an additional available series with a notification of the abnormalities detected and (D) segmentation of the abnormalities overlaid on the scan images.

### Outcome measures

The primary outcome measure was the difference measured in Area Under the Receiver Operator Curve (AUC) of readers in identifying a scan as containing one or more critical findings (intracranial haemorrhage subcategorised into 5 subtypes, hypodensity suggestive of infarct, with and without AI assistance. Secondary outcome measures included differences in reader sensitivity, specificity and confidence with and without AI assistance, the stand-alone diagnostic performance of the qER AI versus ground truth for each target abnormality, and median scan interpretation time.

### Sample size and power calculation

A sample of 30 readers and minimum 135 scans (82 with presence of critical findings and 53 with no critical findings) was estimated to have a minimum 80% power at a type I error rate of 5% to detect a minimum difference in readers’ AUC of 5%, assuming a large inter-reader and intra-reader variability of 0.3 and 0.05, respectively, a 0.35 conservative correlation between readers, and anticipated average readers’ AUC of 0.75, guided by previous literature.^16,17^

### Statistical analyses

The stand-alone performance of qER algorithm was compared with the ground truth generated by the neuroradiologists, using the continuous probability score from the algorithm for the AUC analyses, and binary classification results for the evaluation of sensitivity, specificity, positive predictive value and negative predictive value.

The difference in AUC of readers with and without AI was tested based on the Obuchowski-Rockette model for MRMC analysis which models the data using a two-way mixed effects analysis of variance model treating readers and cases (images) as random effects and effect of AI as a fixed effect with recommended adjustment to df by Hillis et al. Sensitivity and specificity was analysed as part of this model. The main analysis was performed as a single pool including all groups and sites. Prespecified subgroup analyses were performed for the following variables: professional group (radiologist vs ED clinician vs radiographer), post-graduation experience level (junior <5 years, middle grade 5 to 10 years, and senior >10 years), pathological finding and difficulty of image.^16-18^

The median review time per scan with vs without AI was compared using a non-parametric Wilcoxon sign-rank test.

### Patient and public involvement

This study was presented to the Oxford ACUTECare PPI group who supported the study and its aims, and influenced design, data management and dissemination strategies.

## Results

### Baseline characteristics

Baseline characteristics are summarised in Table S1 in supplementary material. Of the 150 images in the dataset, 98 were defined by the radiologist panel as containing one or more critical abnormalities. 30 readers (10 ED clinicians, 10 general radiologists, and 5 radiographers) with in the prespecified seniority classifications each interpreted all 150 cases both with and without AI, totalling 9000 individual interpretations for analysis.

### Algorithm versus ground truth

Retrospective analysis of the diagnostic performance of the qER algorithm versus ground truth is presented in Table S2 in supplementary material, and Figure 3. The algorithm showed strong overall diagnostic performance for all abnormality subgroups with AUCs ranging from 0.821 (95% CI 0.740 - 0.903) to 0.986 (95% CI 0.969 - 1.000), with the exception of mass effect, for which it showed poor discriminative ability in this study with an AUC of 0.604 (95% CI 0.435 - 0.774). Lower sensitivities (<0.80) were seen for extradural haemorrhage (0.692, 95% CI 0.613 - 0.766), intraparenchymal haemorrhage (0.576, 95% CI 0.490 - 0.654), intraventricular haemorrhage 0.650 (95% CI 0.571 - 0.729), mass effect (0.286, 95% CI 0.216 - 0.366) and fracture (0.727, 95% CI 0.648 - 0.796); lower specificities (<0.80) were seen for infarct (0.699, 95% CI 0.620 - 0.772) and mass effect (0.727, 95% CI 0.648 - 0.796).

**Figure 3.**
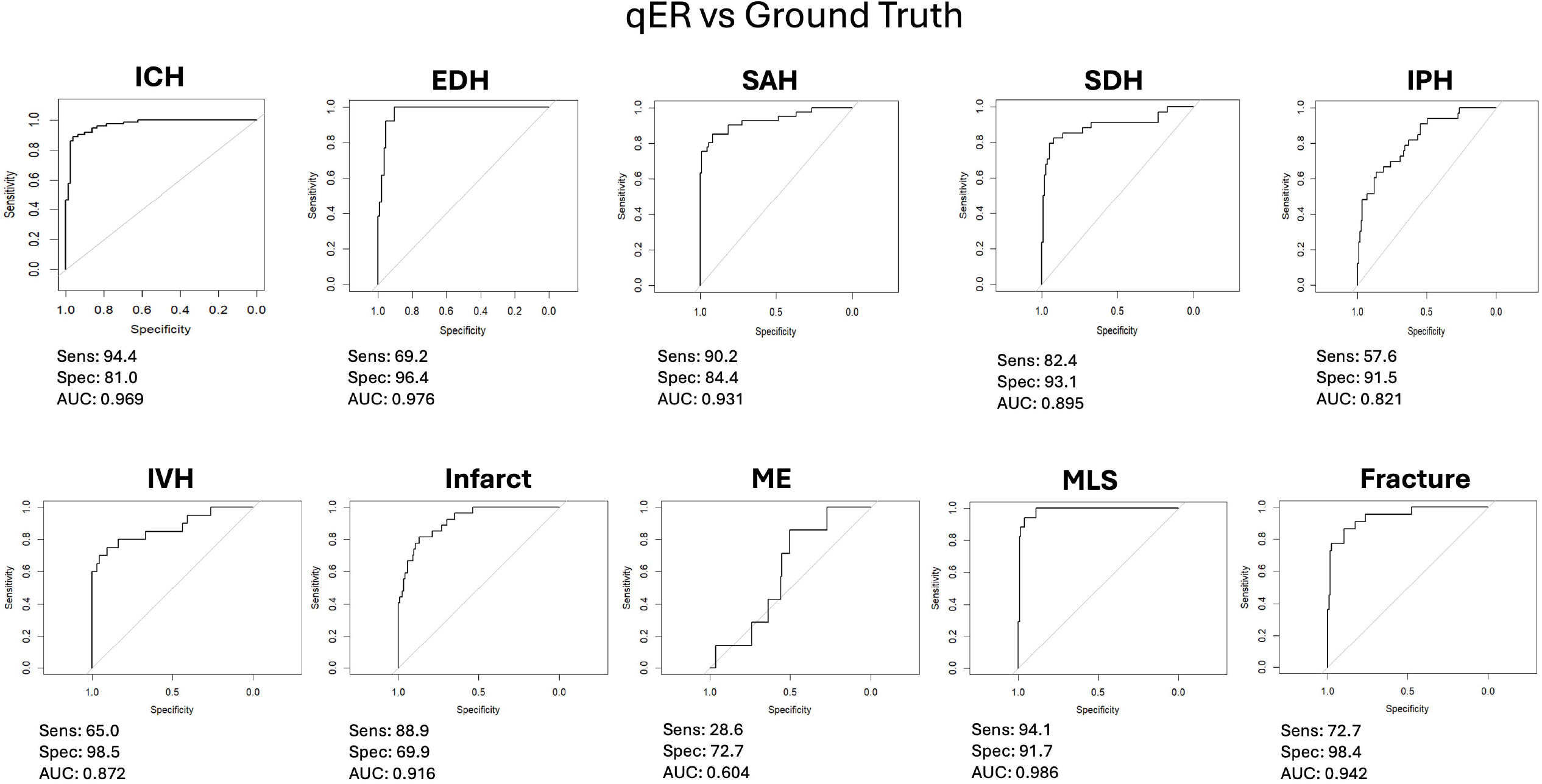
Diagnostic performance of algorithm versus ground truth depicted as area under receiver operator curve (AUC) for different pathological subgroups including Intracranial Haemorrhage (ICH), Extradural Haemorrhage (EDH), Subarachnoid Haemorrhage (SAH), Subdural Haemorrhage (SDH), Intraparenchymal Haemorrhage (IPH), Intraventricular Haemorrhage (IVH), Infarct, Mass Effect (ME), Midline Shift (MLS) and Fracture. Sensitivity (Sens) and specificity (Spec) are given as per the default threshold settings for the algorithm for each pathological subgroup.

### Overall reader performance

Changes in overall (pooled) reader performance for different pathological subgroups are presented in table S3 in supplementary material, and figure 4. Diagnostic performance for the detection of critical abnormality measured by area under the curve showed no statistically significant change, however a statistically significant increase in pooled sensitivity for critical abnormality was observed from 82.8% to 89.8% (difference +7.0% 95% CI 3.4% to 10.6%, p<0.001). This was accompanied however by a corresponding decrease in specificity from 84.5% to 78.9% (difference −5.5%, 95% CI −0.09% to 11.0%, p=0.046). Statistically significant increases were demonstrated for sensitivity across all main pathology subgroups, and for AUC in intracranial haemorrhage (0.853 to 0.956, difference +0.104, 95%CI 0.0602 to 0.147, p<0.001), infarct (0.782 to 0.846, difference +0.0636, 95%CI 0.0211 to 0.106, p=0.0035) and fracture (0.845 to 0.902, difference +0.0571 95%CI 0.0162 to 0.098, p=0.007) subgroups, with a decrease in specificity seen in the midline shift subgroup from 97.4% to 94.2% (difference −3.2%, 95%CI −0.94% to 5.5%, p=0.001)(see Supplementary Materials Tables S5 to S11).

**Figure 4.**
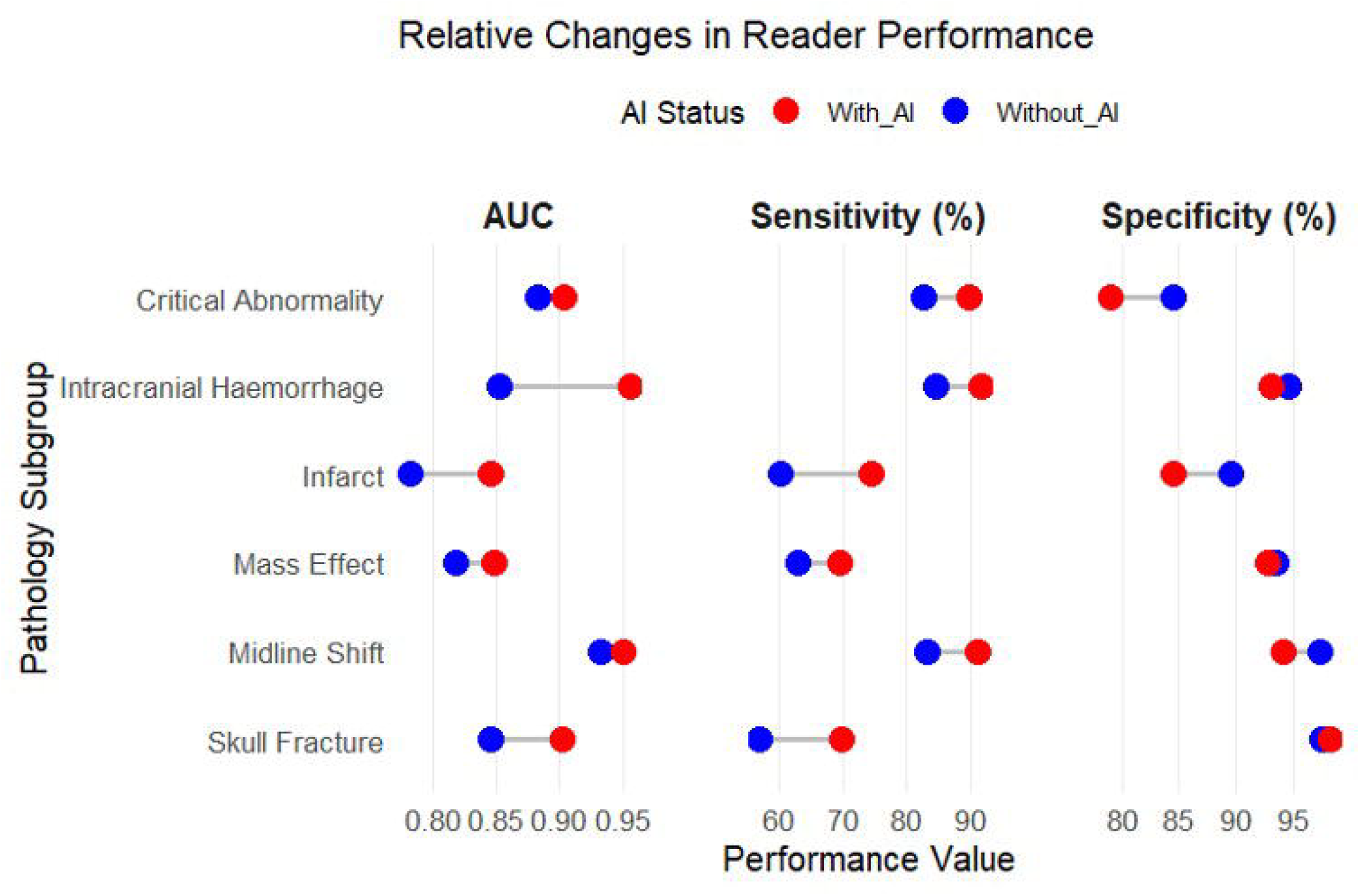
Relative changes in pooled reader performance measured in terms of area under the ROC curve (AUC), sensitivity and specificity for different pathology subgroups

### Subgroup analyses

Relative performance for different reader subgroups in detecting critically abnormal scans are summarised in table S4 in supplementary material, and figure 5. Subgroup analyses for Emergency medicine doctors demonstrated no significant change in AUC but a large increase in sensitivity for the detection of critically abnormal scans (81.0% to 88.0%, difference +6.9%, 95% CI 0.02.8% to 11.1%) p = 0.001), with increases in sensitivity seen across all pathology subgroups (see supplemental materials), and increases in AUC for ICH, infarct and fracture subgroups; however a decrease in specificity was observed in the infarct subgroup. General radiologists showed no significant change in ability to detect critical abnormality with AI, but demonstrated a large increase in AUC and small increase in sensitivity for intracranial haemorrhage from 84.2% (95% CI 75.1% to 93.2%) to 97.8% (95% CI 96.0% to 99.6%, p= 0.007), and 93.9% (95% CI 89.9 to 98.0) to 95.4% (95% CI 91.7% to 99.0%, p = 0.014) respectively (see Supplemental Materials table S6). Radiographers demonstrated increases in the AUC for intracranial haemorrhage, infarct and mass effect subgroups, with the latter two also showing an increase in sensitivity. Senior readers demonstrated a significant increase in sensitivity for critical abnormality (83.3% to 90.5%, difference +7.2, 95% CI 2.3% to 12.1%), otherwise no statistically significant difference in critical abnormality detection was seen across seniority subgroups.

**Figure 5.**
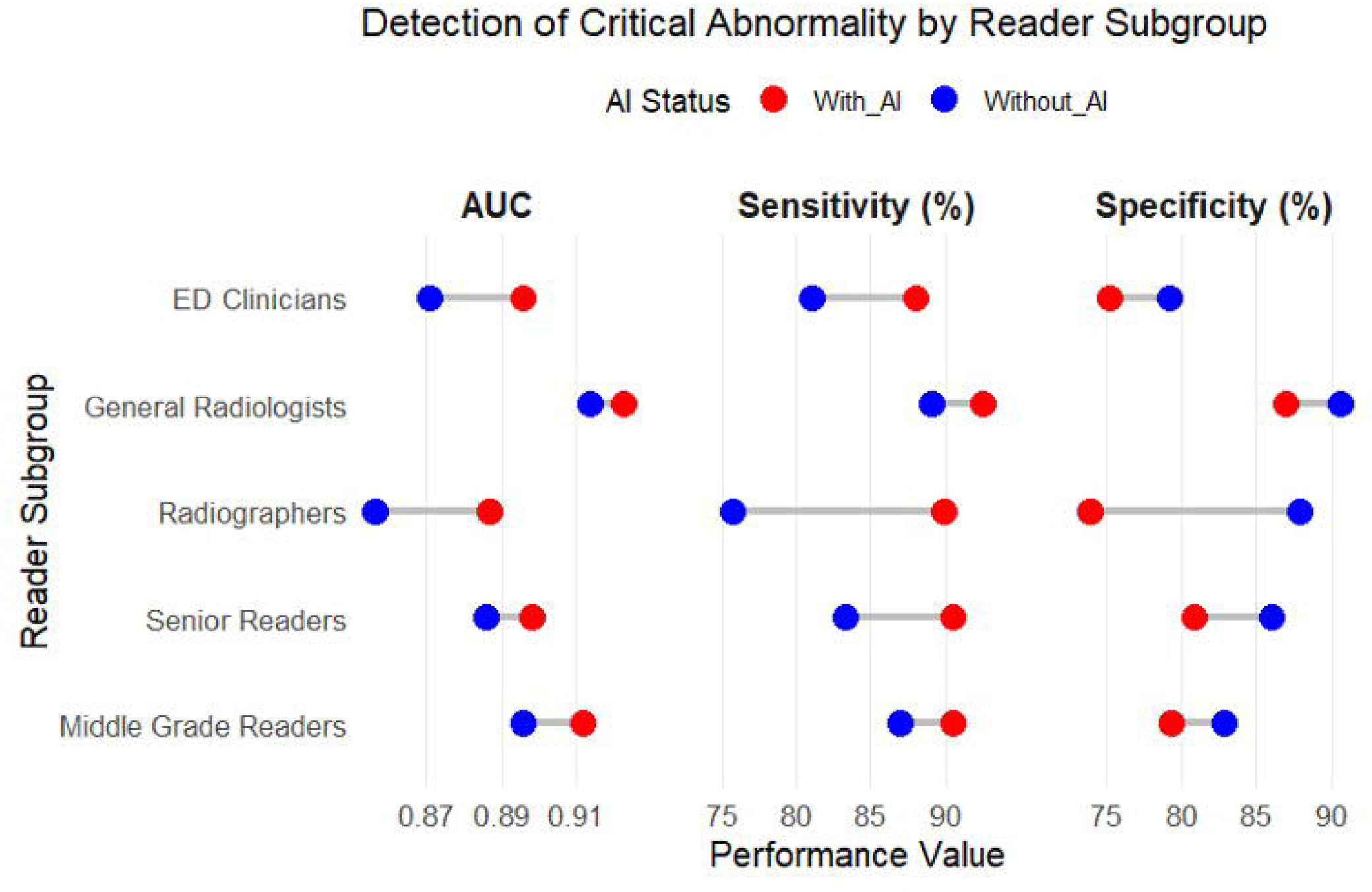
Relative reader subgroup performance for Abnormality and ICH, infarct, mass effect, fracture.

### Interpretation Time

Mean interpretation time was found to be 192 seconds per case in phase 1 (without AI) and reduced to 173 seconds per case in phase 2 (with AI) (p < 0.001). Relevant data were winsorized at a 95% threshold to exclude extreme outliers.

## Discussion

This study evaluated the impact of an AI-assisted image interpretation on the diagnostic performance of a group of radiologists, radiographers and Emergency Medicine clinicians routinely involved in the care of patients undergoing NCCTH. Key findings included: i) a strong overall diagnostic performance of the algorithm measured as AUC against an enhanced ‘ground truth’ reference standard of senior neuroradiologist reporting for 9 out of 10 pathological subgroups, ii) a significant increase in pooled reader sensitivity for the detection of critical abnormality with AI assistance coupled with a comparable decrease in specificity, however no statistically significant change in AUC, and iii) a marked increase in the sensitivity of emergency medicine clinicians for the detection of critical abnormality with AI, comparable to the unaided performance of general radiologists. No difference in effect was seen across different seniority subgroups with AI assistance.

The qER algorithm has previously been retrospectively tested on an external validation data set of 491 NCCTH scans from patients in India. In that study the AUC for ICH, skull fracture, midline shift and mass effect was determined as 0.941 (95% CI: 0.919 to 0.965), 0.962 (95% CI: 0.92 to 1.0), 0.970 (95% CI: 0.94 to 0.999) and 0.922 (95% CI: 0.888 to 0.955), respectively. In a subsequent Swedish stroke registry study, qER was found to have 97% sensitivity in detecting non-traumatic ICH. This study demonstrated similar performance characteristics of in most respects, however low sensitivities of qER were seen for intraparenchymal haemorrhage (0.58), mass effect (0.29), and intraventricular haemorrhage (0.65), and low specificities (<0.80) were seen for infarct (0.70) and mass effect (0.73). This serves as an important reminder of variability in the performance of AI-assisted image interpretation algorithms on a case-by-case basis. Readers in this study showed no improvement in sensitivity for detection of intraparenchymal or intraventricular haemorrhage (see supplementary material table S5), a decrease in sensitivity for midline shift (from a high baseline) and a significant decrease in specificity for infarct detection with AI assistance, suggesting that limitations in algorithm performance can potentially translate to an adverse effect on the performance of human readers in some use cases, which is of critical importance in considering options for real-world deployment, and the need to inform readers re the reliability and relative ‘confidence’ of the algorithm output for a given finding.

There are no prior reports on the impact of qER assistance on readers, hence the AI-REACT study is the first to investigate this effect. Few clinical reader studies have been undertaken for any CT Head interpretation algorithm.^19^ Of note, a 2023 paper by Buchlak et al evaluated the impact of a deep learning algorithm for 22 pathological findings (containing 192 subcategory findings) for CT head on the reporting accuracy of 32 radiologists. Those findings which demonstrated an AUC > 0.80 (n=144, average AUC 0.93) were then incorporated into an MRMC, in which assisted and unassisted radiologists demonstrated an average AUC of 0.79 and 0.73 across 22 grouped parent findings and 0.72 and 0.68 across 189 child findings, respectively. When assisted by the model, radiologist AUC was significantly improved for 91 findings, and reading time was significantly reduced. The average algorithm AUC obtained in that analysis was comparable with those measured for qER in this study, however markedly lower AUC for skilled radiologist readers both with and without AI assistance implies significant differences between both the reader group and datasets, which in the Buchlak study were randomly selected rather than consecutively derived from clinical practice, and hence caution should therefore be exercised in comparing the results.

Whilst several evaluations of AI-assisted image interpretation have indicated a positive impact on radiologist performance, in this study AI assistance showed limited potential to improve even non-specialist radiologists when applied to a dataset which used consecutive cases and was therefore derived more closely from routine clinical practice. This should temper expectations and assumptions regarding the impact of AI-assisted image interpretation in this context, i.e. skilled radiologist reporting, though this finding may reflect a lower number of ‘difficult’ cases in the dataset used in this study, and the removal of factors such as distraction and fatigue which may impair radiologist performance in real world settings. Furthermore, the decreases in pooled reader specificity for detection of critical abnormality and certain pathology subgroups indicate the potential for AI assistance to adversely affect reader performance in some circumstances. Equally, algorithms optimised for sensitivity may support non-specialist readers, but bias may lead to ‘overcall’, which needs to be taken into account when considering potential roles for AI assistance. AI did not significantly improve non-specialist radiographer accuracy to the same levels as the other specialty groups, indicating that a priori reader skill remains important in assisted accuracy, and that AI assistance alone is unlikely to replace experience in this context. Conversely, the lack of difference between seniority subgroups suggests that clinical experience is not necessarily an indicator of skill in interpreting CT head images in the context of evaluations such as this. Assisted image interpretation AI improved the diagnostic performance of ED clinicians to levels comparable with that of unaided general radiologists, who represent a pragmatic benchmark for current radiological clinical practice. This suggests that it may be possible to identify subgroups of patients on a clinical basis for whom the interpretation of ED clinicians may be safe and effective enough to allow clinical actions to be taken prior to the availability of a radiological report. Future studies should evaluate this potential on a prospective clinical basis, and should explore the optimisation of algorithm threshold and calibration to increase negative predictive value facilitate the reliable identification of ‘normal’ scans to facilitate early ED discharge.

This study was designed to reflect current trends in the methodology of assessing clinicians and to facilitate comparison with other studies reporting similar evaluations, and as such chose AUC as a primary outcome measure of reader accuracy, utilising self-reported reader confidence as a variable performance metric. This is useful in demonstrating changes in performance between paired unassisted and assisted groups, though it can be misleading to use this for cross comparison between different reader subgroups, or between readers and the algorithm. Whilst AUC is useful to understand the potential of an AI algorithm to accurately identify pathologies and to determine the optimum operating thresholds for reporting a pathological findings as present or absent, in clinical practice need for a specific pre-determined threshold renders this metric misleading in terms of clinical impact, as algorithms which demonstrate a high AUC overall may still have relatively low sensitivity or specificity at default operating thresholds which may limit clinical applicability, hence the need to report and consider all performance metrics in evaluating these technologies.

### Strengths

This study evaluated the impact of the artificial intelligence (AI) tool on diagnostic accuracy, speed and confidence, in its most realistic use-case, as an assistant to healthcare professionals rather than in isolation. It represents the first UK-based multicentre validation of an AI for non-contrast CT head scans trained on a large data set (300⍰000 head CTs). The dataset constructed used a systematic approach to collect consecutive cases derived from routine clinical datasets, increasing validity and generalisability of results compared to other studies which have used more subjective and less transparent approaches to creating image datasets.

The reader group itself is large (n=30) compared to other multicase multireader studies.^20^ 5 readers represents a typical minimum group size for such studies, so each reader speciality subgroup in our study included at least five readers, allowing for independent subgroup analyses.^17^ Nevertheless, variation in reader performance occurs on an individual basis which may limit the generalisability of findings. The reader group includes non-radiologists (emergency medicine clinicians and radiographers) among the healthcare professionals that may benefit from AI assistance. This allows the potential utility of AI-assisted CT Head interpretation to be explored in use cases other than supporting the diagnostic performance of radiologists

### Limitations

This was an online study using a curated dataset with an artificially high prevalence of abnormal images in the selected scans, which was enriched in order to achieve statistical power to detect the impact of AI assistance. Although necessary to facilitate an important evaluation of diagnostic accuracy, this limits the immediate generalisability of results to real-life clinical performance. Scans with postoperative changes and significant artefacts (eg, patient movement) fall outside the AI’s scope of training and were excluded from the study. Clinical data was limited to the clinical vignette on request form – this reflects real-world practice for radiologists, but clinician readers would have not been able to judge who was high risk for the presence of pathology, e.g. evident clinically significant injury/presentation, which may have informed their diagnostic decisions when interpreting the scans in a real life clinical context.

## Conclusion

Use of AI-assisted image interpretation for NCCTH significantly increased the pooled sensitivity of a group of radiologist and clinician readers in detecting critical abnormalities, however this was accompanied by a comparable decrease in specificity. Subgroup analysis showed limited benefit to skilled radiologist readers, but demonstrated a significant increase in the sensitivity of ED clinicians in detecting abnormality, to a level comparable to that of unaided radiologists. These findings should be fully explored prospectively to validate these results and identify potential use cases for this application.

## Data Availability

The study data will be freely available upon request to the corresponding author.

## Ethics and dissemination

The study has been approved by the UK Health Research Authority (IRAS number 310995, approved 13/12/2022). The use of anonymised retrospective CT scans has been authorised by the Caldicott Guardian and information governance team at Oxford University Hospitals NHS Foundation Trust. Readers will provide written informed consent and will be able to withdraw at any time.

The study is registered at Clinicaltrials.gov (NCT06018545), and the ISRCTN registry (ISRCTN17560291). The results of the study will be presented at relevant conferences and published in peer-reviewed journals. The detailed study protocol will be freely available upon request to the corresponding author. Further dissemination strategy will be strongly guided by our PPIE activities. This will be based on co-productions between patient partners and academics and will involve media pieces (mainstream and social media) as well as communication through charity partners.

## Authors’ contributions

AN and SA led the design of the project, with contributions from RS, ATEM, DR, SK, SG, JO, KB, AR, TD, MTG, MN, RD, MH, KV, JG, NW, NS, AC, FK, DL, and HS. SA, RS, FK and NS and AC and reviewed the image dataset. KB and AR are the primary ground-truthers, with arbitration from TD. NS manages the online CT reading platform and assisted in data collection and management. ATEM registered the study and coordinates reader recruitment and data collection. AN, and SA wrote the manuscript. NW and JG led the PPI activities.

