## supplementary material for "Artificial Intelligence-assisted reader evaluation in acute CT head interpretation (AI-REACT): a multireader multicase study"

**AI-REACT Reader Study Group (Group authorship):**

Satish Golla^3^, Irfan Ullah Akbarkhan^8^, Laura Hunter^8^, Nazreen Kaneez^8^, Ana Nicolescu^8^, Emma Kelliher^8^, Kyle Stephenson^8^, Shair Ali^8^, Danielle Benson^8^, Fiona Hunter^9^, Ross Hunter^9^, Benjamin Scally^9^, Rhys Worgan^9^, Louise Hartley^9^, Ryan Grech^9^, Martine Walker^9^, Neil Mitchell^9^, Ravi Shashikala^6^, Alice Gibson^6^, Hélène Matte^6^, Shubhendu Kulshrestha^6^, Hannah Yang^6^, Radoslaw Rippel^6^, Roland Amoah^6^, Zahi Qamhawi^6^, Thomas Millard^6^, Avneet Gill^6^, Michael Thompson^10^, Josh Beck^10^, Harsh Merchant^10^, Ben Lockwood^10^, Nabeeha Salik^17^

**Affiliations:**

^1^ Oxford Clinical Artificial Intelligence Research (OxCAIR), Oxford University Hospitals NHS Foundation Trust

^2^ Emergency Medicine Research Oxford, Oxford University Hospitals NHS Foundation Trust

^3^ Qure.ai, Bangalore, India

^4^ Qure.ai Technologies Limited, London, UK

^5^ Department of Primary Health Care Sciences, University of Oxford, Oxford, UK

^6^ Oxford University Hospitals NHS Foundation Trust, Oxford, UK

^7^ Department of Clinical Radiology, Cambridge University Hospitals NHS Foundation Trust, Cambridge, UK

^8^ Guy's & St Thomas' NHS Foundation Trust, London, UK

^9^ NHS Greater Glasgow and Clyde, Glasgow, UK

^10^ Northumbria Healthcare NHS Foundation Trust, Newcastle upon Tyne, UK

^11^ School of Biomedical Science, King’s College London, London UK

^12^ Emergency Department, Northumbria Specialist Emergency Care Hospital, Cramlington, UK

^13^ Clinical Scientific Computing, Guy’s and St Thomas’ NHS Foundation Trust, London, UK

^14^ College of Health, Psychology & Social Care, University of Derby, Derby, UK

^15^ University College London NHS Foundation Trust, London, UK

^16^ School of Allied and Public Health Professions, Canterbury Christ Church University, Canterbury, UK

^17^ RAIQC, LTD, Oxford, UK

^#^ First Author

Corresponding Author:

Dr Alex Novak

RCEM Associate Professor of Emergency Medicine

Co-Director Oxford Clinical Artificial Intelligence Research (OxCAIR)

Director of Emergency Medicine Research Oxford (EMROx)

Consultant in Emergency Medicine and Ambulatory Care
Oxford University Hospitals NHS Foundation Trust

**Supplementary Figures**

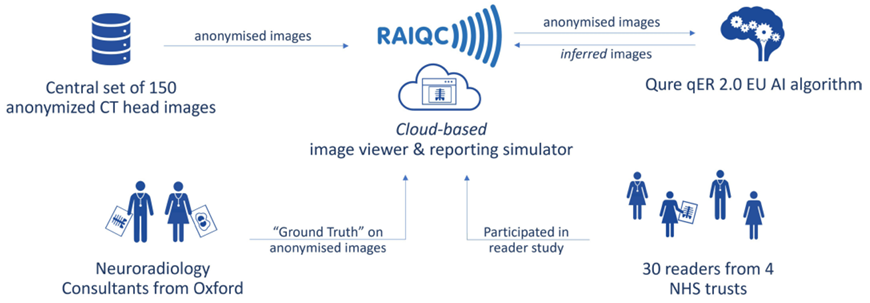

Figure S1. AI-REACT dataflow diagram. Scans selected for the study were anonymised in accordance with Oxford University Hospitals NHS Foundation Trust information governance protocol using the Insignia Insight Anonymisation tool and uploaded to the secure image viewing platform (www.raiqc.com). Access to the scans was controlled via the study platform using separate user accounts for each reader. The anonymised images were sent securely to Qure servers for inferencing, and outputs transferred back to RAIQC. Data about the readers’ seniority level and professional group was retained to allow group comparisons.

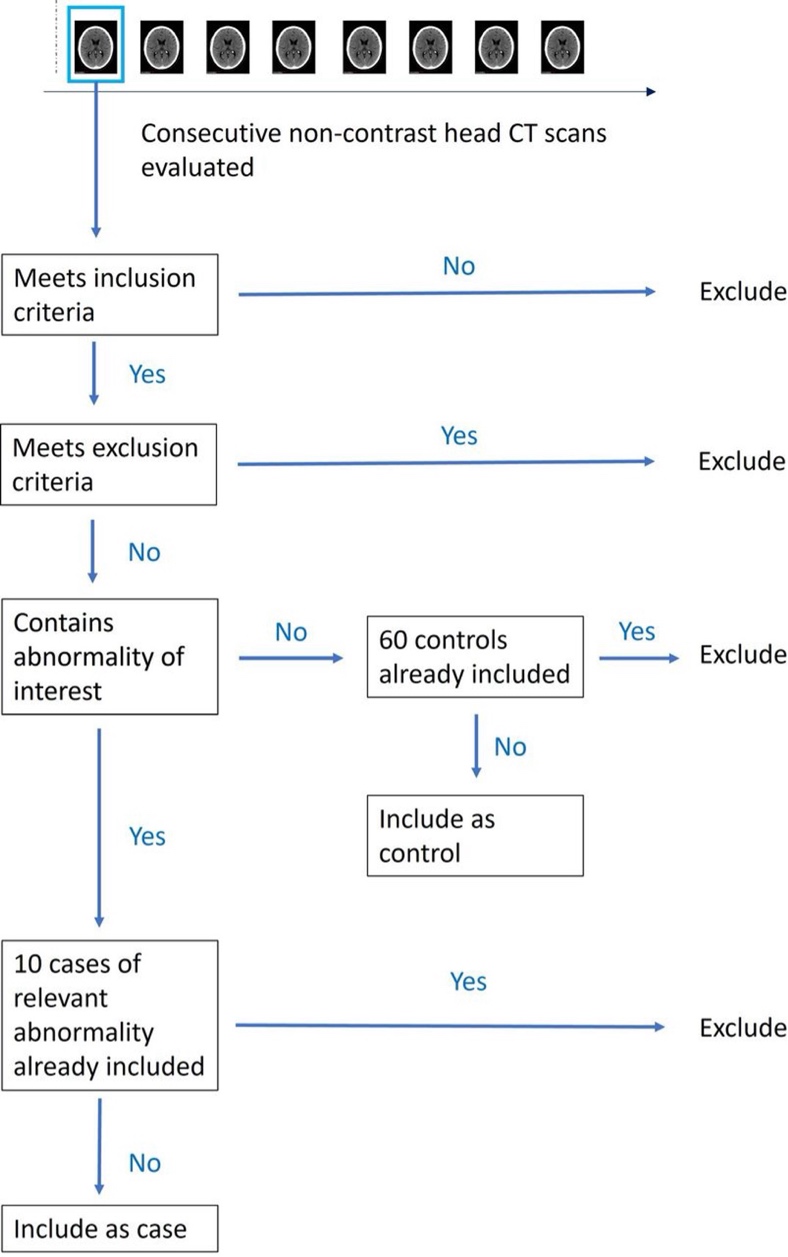

Figure S2. Flow diagram showing AI-REACT Case selection.

**Supplementary Tables**

| **Pathological Finding** | **Number in dataset (n =)** |
| --- | --- |
| No Critical Abnormality | 52 |
| Critical Abnormality | 98 |
| - Intracranial Haemorrhage | 71 |
| - Subdural Haemorrhage | 34 |
| - Subarachnoid Haemorrhage | 41 |
| - Extradural Haemorrhage | 13 |
| - Intraparenchymal Haemorrhage | 33 |
| - Intraventricular Haemorrhage | 20 |
| - Infarct | 27 |
| - Mass Effect | 48 |
| - Midline Shift | 17 |
| - Skull Fracture | 22 |

Table S1. Baseline characteristics of dataset following expert radiology panel review (ground truth)

| Abnormality | AUC (95% CI) | Sensitivity (95% CI) % | Specificity (95% CI) % |
| --- | --- | --- | --- |
| Intracranial Haemorrhage | 0.969 (0.946 - 0.993) | 94.4 (89.8 - 97.7) | 81.0 (74.2 - 87.2) |
| Extradural Haemorrhage | 0.976 (0.954 - 0.999) | 69.2 (61.3 – 76.6) | 96.4 (92.4 – 98.9) |
| Subarachnoid Haemorrhage | 0.931 (0.877 - 0.985) | 90.2 (84.0 – 94.3) | 84.4 (77.9 – 90.0) |
| Subdural Haemorrhage | 0.895 (0.817 - 0.974) | 82.4 (75.6 – 88.4) | 93.1 (88.1 – 96.8) |
| Intraparenchymal Haemorrhage | 0.821 (0.740 - 0.903) | 57.6 (49.0 – 65.4) | 91.5 (85.6 – 95.3) |
| Intraventricular Haemorrhage | 0.872 (0.768 - 0.976) | 65.0 (57.1 – 72.9) | 98.5 (95.3 – 99.8) |
| Infarct | 0.916 (0.863 - 0.969) | 88.9 (82.5 – 93.3) | 69.9 (62.0 – 77.2) |
| Mass Effect | 0.604 (0.435 - 0.774) | 28.6 (21.6 – 36.6) | 72.7 (64.8 – 79.6) |
| Midline Shift | 0.986 (0.969 - 1.000) | 94.1 (88.9 – 97.2) | 91.7 (86.4 – 95.8) |
| Fracture | 0.942 (0.889 - 0.996) | 72.7 (64.8 – 79.6) | 98.4 (95.3 – 99.8) |

Table S2. Summary of diagnostic performance characteristics for the qER 2.0 EU algorithm compared against the reference standard

| **Analysis** | | **Without AI (95% CI)** | **With AI (95% CI)** | **Difference (95% CI)** | **p value** |
| --- | --- | --- | --- | --- | --- |
| **Critical Abnormality** | |  |  |  |  |
| AUC | | 0.883 (0.847, 0.918) | 0.904 (0.872, 0.935) | +0.021 (-0.006, 0.05) | 0.13 |
| Sensitivity (%) | | 82.8 (77.1, 88.5) | 89.8 (85.7, 93.9) | +7.0 (3.3, 10.6) | <0.001 |
| Specificity (%) | | 84.5 (79.5, 89.4) | 78.9 (71.9, 85.9) | -5.5 (-11.0, -0.09) | 0.046 |
| **Intracranial Haemorrhage** | |  |  |  |  |
| AUC | | 0.853 (0.808, 0.898) | 0.956 (0.933, 0.980) | +0.104 (0.060, 0.147) | <0.001 |
| Sensitivity (%) | | 84.6 (78.3, 91.0) | 91.6 (87.1, 96.2) | +7.0 (3.2, 10.9) | <0.001 |
| Specificity (%) | | 94.6 (92.3, 96.9) | 93.1 (89.4, 96.7) | -1.5 (1.2, -4.1) | 0.27 |
| **Infarct** | |  |  |  |  |
| AUC | | 0.782 (0.716, 0.849) | 0.846 (0.790, 0.902) | +0.0636 (0.0211, 0.106) | 0.0035 |
| Sensitivity (%) | | 60.1 (47.9, 72.4) | 74.5 (64.1, 85.0) | +14.4 (6.5, 22.3) | <0.001 |
| Specificity (%) | | 89.6 (85.8, 93.5) | 84.5 (79.2, 89.7) | -5.1 (-1.9, -8.4) | 0.0024 |
| **Mass Effect** | |  |  |  |  |
| AUC | | 0.818 (0.766, 0.870) | 0.848 (0.798, 0.897) | +0.0299 (-0.00111, 0.0609,) | 0.059 |
| Sensitivity (%) | | 63.0 (52.0, 73.9) | 69.6 (59.3, 79.8) | +6.6 (0.96, 12.2) | 0.023 |
| Specificity (%) | | 93.5 (90.6, 96.4) | 92.7 (89.0, 96.4) | -0.8 (-2.9, 1.2) | 0.43 |
|  | **Midline Shift** |  |  |  |  |
|  | AUC | 0.933 (0.885, 0.981) | 0.950 (0.908, 0.992) | 0.0171 (0.0501, -0.0159) | 0.31 |
|  | Sensitivity | 83.3 (71.2, 95.5) | 91.2 (82.2, 100) | +7.8 (0.4, 15.2) | 0.038 |
|  | Specificity | 97.4 (95.8, 99.0) | 94.2 (91.1, 97.2) | -3.2 (-0.94, -5.5) | 0.006 |
| **Skull Fracture** | |  |  |  |  |
| AUC | | 0.845 (0.790, 0.899) | 0.902 (0.847, 0.956) | +0.0571 (0.0162, 0.098) | 0.007 |
| Sensitivity (%) | | 57.0 (43.5, 70.4) | 69.8 (56.1, 83.6) | +12.9 (4.25, 21.51) | 0.0039 |
| Specificity (%) | | 97.6 (96.4, 98.7) | 98.3 (97.0, 99.6) | +0.76 (-0.23,1.739) | 0.13 |

Table S3. Pooled reader analyses for pathological subgroups including Area Under the Receiver Operator Curve (AUC), Sensitivity and Specificity.

| **Reader Subgroup** | **Without AI (95% CI)** | **With AI (95% CI)** | **Difference (95% CI)** | | **p value** |
| --- | --- | --- | --- | --- | --- |
| **ED Clinicians** |  |  |  |  | |
| AUC | 0.871 (0.830, 0.912) | 0.896 (0.859, 0.934) | +0.0255 (-0.014, 0.065) | 0.2 | |
| Sensitivity (%) | 81.0 (75.3, 86.8) | 88.0 (82.8, 93.2) | +0.07 (2.8, 11.1) | 0.001 | |
| Specificity (%) | 79.2 (72.0, 86.5) | 75.2 (65.2, 85.2) | -0.04 (-12.0, 4.0) | 0.31 | |
| **General Radiologists** |  |  |  |  | |
| AUC | 0.914 (0.877, 0.951) | 0.923 (0.892, 0.955) | +0.0093 (0.0348, -0.0162) | 0.44 | |
| Sensitivity (%) | 89.1 (82.5, 95.6) | 92.5 (87.7, 97.2) | +3.4 (-0.4, 7.2) | 0.077 | |
| Specificity (%) | 90.6 (85.2, 95.9) | 87.0 (80.8, 93.1) | -3.6 (2.0, -9.2) | 0.18 | |
| **Radiographers** |  |  |  |  | |
| AUC | 0.856 (0.778, 0.934) | 0.887 (0.833, 0.940) | +0.0306 (0.1014, -0.0402) | 0.34 | |
| Sensitivity (%) | 75.7 (58.5, 92.9) | 89.9 (84.2, 95.6) | +14.2 (30.2, -1.7) | 0.07 | |
| Specificity (%) | 87.9 (81.5, 94.3) | 74.0 (64.0, 83.9) | -14.0 (-0.61, -27.3) | 0.043 | |
| **Senior Readers** |  |  |  |  | |
| AUC | 0.886 (0.851, 0.920) | 0.898 (0.862, 0.934) | +0.0123 (-0.0259, 0.0505) | 0.5 | |
| Sensitivity (%) | 83.3 (76.2, 90.4) | 90.5 (85.5, 95.5) | +7.2 (2.3, 12.1) | 0.007 | |
| Specificity (%) | 86.0 (78.3, 93.8) | 80.9 (72.4, 89.5) | -5.1 (-10.8, 0.61) | 0.078 | |
| **Middle Grade Readers** |  |  |  |  | |
| AUC | 0.896 (0.845, 0.946) | 0.912 (0.876, 0.948) | +0.0165 (0.0561, -0.0232) | 0.39 | |
| Sensitivity (%) | 86.9 (80.6, 93.2) | 90.5 (84.7, 96.4) | +3.6 (-0.17, 7.4) | 0.06 | |
| Specificity (%) | 82.8 (72.6, 93.1) | 79.4 (66.0, 92.9) | -3.4 (7.6, -14.4) | 0.5 | |

Table S4 Detection of critical abnormality by specialty and seniority reader subgroup

| Analysis | Without AI (95% CI) | With AI (95% CI) | Difference (95% CI) | p value |
| --- | --- | --- | --- | --- |
| Extradural |  |  |  |  |
| AUC | 0.854 (0.773, 0.935) | 0.881 (0.816, 0.945) | 0.0269 (0.0748, -0.0211) | 0.27 |
| Sensitivity | 0.638 (0.480, 0.797) | 0.685 (0.534, 0.835) | 0.0462 (0.127, -0.0346) | 0.26 |
| Specificity | 0.972 (0.954, 0.989) | 0.975 (0.956, 0.995) | 0.00389 (0.0117, -0.0039) | 0.33 |
| Subdural |  |  |  |  |
| AUC | 0.777 (0.718, 0.835) | 0.868 (0.819, 0.917) | 0.0913 (0.138, 0.0444) | <0.001 |
| Sensitivity | 0.481 (0.365, 0.596) | 0.678 (0.571, 0.786) | 0.198 (0.277, 0.118) | <0.001 |
| Specificity | 0.969 (0.954, 0.984) | 0.959 (0.937, 0.982) | -0.00948 (0.00762, -0.0266) | 0.28 |
| Subarachnoid |  |  |  |  |
| AUC | 0.809 (0.751, 0.868) | 0.920 (0.889, 0.952) | 0.111 (0.156, 0.0663) | <0.001 |
| Sensitivity | 0.575 (0.454, 0.696) | 0.844 (0.778, 0.909) | 0.268 (0.356, 0.181) | <0.001 |
| Specificity | 0.948 (0.927, 0.969) | 0.904 (0.866, 0.943) | -0.0438 (-0.0179, -0.0698) | 0.001 |
| Intraparenchymal |  |  |  |  |
| AUC | 0.828 (0.768, 0.889) | 0.841 (0.781, 0.900) | 0.0124 (0.0546, -0.0297) | 0.56 |
| Sensitivity | 0.681 (0.570, 0.792) | 0.652 (0.530, 0.775) | -0.0286 (0.0507, -0.108) | 0.48 |
| Specificity | 0.932 (0.902, 0.962) | 0.952 (0.927, 0.977) | 0.0197 (0.0459, -0.00651) | 0.14 |
| Intraventricular |  |  |  |  |
| AUC | 0.803 (0.729, 0.877) | 0.828 (0.749, 0.907) | 0.0247 (0.0722, -0.0229) | 0.31 |
| Sensitivity | 0.550 (0.394, 0.706) | 0.597 (0.432, 0.761) | 0.0467 (0.119, -0.0257) | 0.21 |
| Specificity | 0.987 (0.980, 0.994) | 0.984 (0.970, 0.997) | -0.00333 (0.00672, -0.01338) | 0.51 |

Table S5 Pooled reader performance for intracranial haemorrhage subtypes

| Analysis | Without AI (95% CI) | With AI (95% CI) | Difference (95% CI) | p value |
| --- | --- | --- | --- | --- |
| Abnormality |  |  |  |  |
| AUC | 0.914 (0.877, 0.951) | 0.923 (0.892, 0.955) | 0.0093 (0.0348, -0.0162) | 0.439 |
| Sensitivity | 0.891 (0.825, 0.956) | 0.925 (0.877, 0.972) | 0.034 (0.0724, -0.00438) | 0.0769 |
| Specificity | 0.906 (0.852, 0.959) | 0.870 (0.808, 0.931) | -0.0358 (0.0201, -0.0918) | 0.181 |
| Intracranial Haemorrhage |  |  |  |  |
| AUC | 0.842 (0.751, 0.932) | 0.978 (0.960, 0.996) | 0.136 (0.2268, 0.0455) | 0.007 |
| Sensitivity | 0.939 (0.899, 0.980) | 0.954 (0.917, 0.990) | 0.0141 (0.027275, 0.000894) | 0.038 |
| Specificity | 0.973 (0.949, 0.997) | 0.978 (0.958, 0.999) | 0.00506 (0.01569, -0.00557) | 0.309 |
| Extradural |  |  |  |  |
| AUC | 0.921 (0.853, 0.988) | 0.926 (0.886, 0.966) | 0.00547 (0.0472, -0.0362) | 0.791 |
| Sensitivity | 0.738 (0.558, 0.919) | 0.831 (0.739, 0.923) | 0.0923 (0.2146, -0.0299) | 0.128 |
| Specificity | 0.981 (0.965, 0.997) | 0.976 (0.953, 0.999) | -0.00511 (0.00584, -0.01606) | 0.341 |
| Subdural |  |  |  |  |
| AUC | 0.875 (0.810, 0.939) | 0.889 (0.827, 0.951) | 0.0143 (0.0597, -0.0312) | 0.496 |
| Sensitivity | 0.698 (0.576, 0.819) | 0.724 (0.592, 0.855) | 0.0259 (0.0878, -0.036) | 0.386 |
| Specificity | 0.955 (0.929, 0.982) | 0.963 (0.938, 0.987) | 0.00778 (0.0218, -0.00624) | 0.251 |
| Subarachnoid |  |  |  |  |
| AUC | 0.950 (0.915, 0.984) | 0.964 (0.938, 0.990) | 0.0146 (0.03053, -0.00128) | 0.0692 |
| Sensitivity | 0.885 (0.806, 0.963) | 0.933 (0.873, 0.994) | 0.0487 (0.09082, 0.00662) | 0.0254 |
| Specificity | 0.967 (0.942, 0.992) | 0.953 (0.922, 0.984) | -0.0135 (0.00994, -0.03697) | 0.238 |
| Intraparenchymal |  |  |  |  |
| AUC | 0.907 (0.858, 0.957) | 0.873 (0.813, 0.933) | -0.0342 (0.00122, -0.06972) | 0.0576 |
| Sensitivity | 0.817 (0.715, 0.919) | 0.723 (0.601, 0.844) | -0.0943 (-0.0465, -0.1421) | <0.001 |
| Specificity | 0.958 (0.927, 0.990) | 0.968 (0.947, 0.989) | 0.00957 (0.0315, -0.0124) | 0.371 |
| Intraventricular |  |  |  |  |
| AUC | 0.905 (0.841, 0.970) | 0.876 (0.799, 0.953) | -0.0292 (0.0287, -0.0871) | 0.316 |
| Sensitivity | 0.760 (0.620, 0.900) | 0.735 (0.582, 0.888) | -0.025 (0.0745, -0.1245) | 0.618 |
| Specificity | 0.992 (0.983, 1.000) | 0.992 (0.984, 1.000) | 0.000769 (0.01067, -0.00913) | 0.871 |
| Infarct |  |  |  |  |
| AUC | 0.850 (0.765, 0.936) | 0.874 (0.812, 0.936) | 0.0234 (0.0732, -0.0264) | 0.316 |
| Sensitivity | 0.720 (0.568, 0.872) | 0.792 (0.674, 0.910) | 0.072 (0.1541, -0.0101) | 0.0806 |
| Specificity | 0.948 (0.921, 0.975) | 0.934 (0.897, 0.970) | -0.0144 (0.0184, -0.0472) | 0.349 |
| Mass Effect |  |  |  |  |
| AUC | 0.888 (0.846, 0.930) | 0.875 (0.820, 0.929) | -0.0135 (0.017, -0.044) | 0.344 |
| Sensitivity | 0.827 (0.731, 0.923) | 0.812 (0.689, 0.936) | -0.0146 (0.0385, -0.0676) | 0.55 |
| Specificity | 0.891 (0.827, 0.955) | 0.881 (0.809, 0.954) | -0.0098 (0.027, -0.0466) | 0.562 |
| Midline Shift |  |  |  |  |
| AUC | 0.953 (0.908, 0.999) | 0.951 (0.907, 0.994) | -0.00265 (0.0234, -0.0287) | 0.823 |
| Sensitivity | 0.900 (0.790, 1.000) | 0.912 (0.829, 0.995) | 0.0118 (0.077, -0.0534) | 0.693 |
| Specificity | 0.973 (0.953, 0.993) | 0.962 (0.935, 0.989) | -0.0113 (0.00444, -0.027) | 0.146 |
| Skull Fracture |  |  |  |  |
| AUC | 0.933 (0.884, 0.981) | 0.929 (0.879, 0.978) | -0.00391 (0.0199, -0.0277) | 0.73 |
| Sensitivity | 0.832 (0.717, 0.947) | 0.836 (0.723, 0.950) | 0.00455 (0.0369, -0.0278) | 0.758 |
| Specificity | 0.979 (0.962, 0.995) | 0.981 (0.963, 1.000) | 0.00234 (0.01255, -0.00786) | 0.626 |

Table S6: General Radiology subgroup

| **Analysis** | **Without_AI** | **With_AI** | **Difference** | **p value** |
| --- | --- | --- | --- | --- |
| Abnormality AUC | 0.871 (0.830, 0.912) | 0.896 (0.859, 0.934) | 0.0255 (0.065, -0.014) | 0.199 |
| Abnormality Sens | 0.81 (0.753, 0.868) | 0.88 (0.828, 0.932) | 0.0694 (0.1107, 0.0282) | 0.00132 |
| Abnormality Spec | 0.792 (0.720, 0.865) | 0.752 (0.652, 0.852) | -0.0403 (0.0398, -0.1203) | 0.31 |
| ICH AUC | 0.880 (0.836, 0.923) | 0.948 (0.921, 0.976) | 0.0686 (0.1073, 0.0299) | <0.001 |
| ICH Sens | 0.805 (0.731, 0.878) | 0.894 (0.838, 0.949) | 0.0892 (0.1398, 0.0386) | <0.001 |
| ICH Spec | 0.934 (0.900, 0.968) | 0.917 (0.869, 0.966) | -0.0169 (0.0238, -0.0575) | 0.406 |
| EDH AUC | 0.827 (0.725, 0.928) | 0.851 (0.761, 0.941) | 0.0242 (0.089, -0.0406) | 0.461 |
| EDH Sens | 0.605 (0.422, 0.788) | 0.595 (0.389, 0.801) | -0.0103 (0.104, -0.124) | 0.859 |
| EDH Spec | 0.975 (0.957, 0.992) | 0.980 (0.962, 0.998) | 0.00535 (0.01424, -0.00353) | 0.228 |
| SDH AUC | 0.741 (0.672, 0.811) | 0.858 (0.796, 0.920) | 0.116 (0.1775, 0.0554) | <0.001 |
| SDH Sens | 0.375 (0.246, 0.503) | 0.655 (0.527, 0.783) | 0.28 (0.379, 0.182) | <0.001 |
| SDH Spec | 0.984 (0.974, 0.995) | 0.962 (0.939, 0.985) | -0.0224 (-0.000112, -0.044716) | 0.0489 |
| SAH AUC | 0.762 (0.695, 0.829) | 0.898 (0.861, 0.935) | 0.136 (0.1947, 0.0775) | <0.001 |
| SAH Sens | 0.436 (0.299, 0.572) | 0.783 (0.697, 0.868) | 0.347 (0.452, 0.242) | <0.001 |
| SAH Spec | 0.936 (0.907, 0.966) | 0.883 (0.832, 0.935) | -0.0529 (-0.0191, -0.0866) | 0.0025 |
| IPH AUC | 0.816 (0.747, 0.885) | 0.845 (0.783, 0.906) | 0.0287 (0.0876, -0.0302) | 0.337 |
| IPH Sens | 0.663 (0.541, 0.785) | 0.640 (0.508, 0.772) | -0.0229 (0.0853, -0.131) | 0.674 |
| IPH Spec | 0.907 (0.869, 0.946) | 0.941 (0.908, 0.974) | 0.0336 (0.07306, -0.00581) | 0.0928 |
| IVH AUC | 0.759 (0.670, 0.848) | 0.803 (0.713, 0.894) | 0.0443 (0.1158, -0.0272) | 0.22 |
| IVH Sens | 0.447 (0.271, 0.623) | 0.520 (0.339, 0.701) | 0.0733 (0.15636, -0.00969) | 0.0823 |
| IVH Spec | 0.985 (0.973, 0.996) | 0.982 (0.966, 0.998) | -0.00256 (0.00917, -0.0143) | 0.666 |
| Infarct AUC | 0.760 (0.683, 0.836) | 0.835 (0.768, 0.902) | 0.0751 (0.1399, 0.0104) | 0.0234 |
| Infarct Sens | 0.549 (0.412, 0.687) | 0.704 (0.571, 0.837) | 0.155 (0.2644, 0.0449) | 0.00629 |
| Infarct Spec | 0.877 (0.825, 0.929) | 0.817 (0.748, 0.885) | -0.0608 (-0.0204, -0.1012) | 0.00423 |
| ME AUC | 0.810 (0.752, 0.868) | 0.847 (0.791, 0.903) | 0.0372 (0.07935, -0.00491) | 0.0814 |
| ME Sens | 0.590 (0.474, 0.706) | 0.657 (0.543, 0.771) | 0.0667 (0.1323, 0.00103) | 0.0468 |
| ME Spec | 0.950 (0.927, 0.974) | 0.946 (0.913, 0.978) | -0.00458 (0.0241, -0.0333) | 0.753 |
| MLS AUC | 0.934 (0.883, 0.985) | 0.962 (0.926, 0.997) | 0.0279 (0.0669, -0.011) | 0.157 |
| MLS Sens | 0.831 (0.697, 0.965) | 0.929 (0.831, 1.000) | 0.098 (0.19294, 0.00314) | 0.043 |
| MLS Spec | 0.978 (0.962, 0.994) | 0.937 (0.903, 0.972) | -0.0411 (-0.0132, -0.069) | 0.00422 |
| Fracture AUC | 0.811 (0.745, 0.877) | 0.887 (0.824, 0.949) | 0.0758 (0.1398, 0.0118) | 0.0218 |
| Fracture Sens | 0.455 (0.300, 0.609) | 0.615 (0.458, 0.772) | 0.161 (0.2767, 0.0445) | 0.00792 |
| Fracture Spec | 0.971 (0.955, 0.986) | 0.982 (0.967, 0.997) | 0.0109 (0.0257, -0.00383) | 0.146 |

Table S7: ED subgroup

| **Analysis** | **Without_AI** | **With_AI** | **Difference** | **p value** |
| --- | --- | --- | --- | --- |
| Abnormality AUC | 0.856 (0.778, 0.934) | 0.887 (0.833, 0.940) | 0.0306 (0.1014, -0.0402) | 0.342 |
| Abnormality Sens | 0.757 (0.585, 0.929) | 0.899 (0.842, 0.956) | 0.142 (0.3015, -0.0169) | 0.0697 |
| Abnormality Spec | 0.879 (0.815, 0.943) | 0.740 (0.640, 0.839) | -0.14 (-0.00613, -0.27312) | 0.0426 |
| ICH AUC | 0.793 (0.715, 0.871) | 0.936 (0.903, 0.969) | 0.143 (0.2256, 0.0602) | 0.00305 |
| ICH Sens | 0.783 (0.594, 0.972) | 0.910 (0.851, 0.969) | 0.127 (0.283, -0.0295) | 0.089 |
| ICH Spec | 0.924 (0.891, 0.957) | 0.876 (0.789, 0.963) | -0.0481 (0.0298, -0.126) | 0.186 |
| EDH AUC | 0.802 (0.732, 0.872) | 0.880 (0.792, 0.967) | 0.0775 (0.1723, -0.0172) | 0.0955 |
| EDH Sens | 0.538 (0.357, 0.720) | 0.662 (0.466, 0.857) | 0.123 (0.321, -0.075) | 0.16 |
| EDH Spec | 0.943 (0.905, 0.981) | 0.961 (0.923, 0.998) | 0.0175 (0.03479, 0.000247) | 0.0469 |
| SDH AUC | 0.685 (0.532, 0.839) | 0.855 (0.779, 0.932) | 0.17 (0.3122, 0.0283) | 0.0267 |
| SDH Sens | 0.365 (0.168, 0.562) | 0.659 (0.516, 0.801) | 0.294 (0.459, 0.13) | 0.00235 |
| SDH Spec | 0.950 (0.892, 1.000) | 0.945 (0.911, 0.979) | -0.00517 (0.0287, -0.0391) | 0.746 |
| SAH AUC | 0.671 (0.503, 0.838) | 0.900 (0.841, 0.958) | 0.229 (0.3602, 0.0972) | 0.00838 |
| SAH Sens | 0.374 (0.064, 0.685) | 0.846 (0.720, 0.973) | 0.472 (0.713, 0.231) | 0.00444 |
| SAH Spec | 0.946 (0.890, 1.000) | 0.868 (0.803, 0.934) | -0.0775 (-0.0156, -0.1394) | 0.0187 |
| IPH AUC | 0.707 (0.592, 0.821) | 0.764 (0.687, 0.840) | 0.0571 (0.1445, -0.0304) | 0.175 |
| IPH Sens | 0.463 (0.184, 0.741) | 0.549 (0.411, 0.686) | 0.0857 (0.31, -0.139) | 0.377 |
| IPH Spec | 0.955 (0.915, 0.995) | 0.953 (0.921, 0.985) | -0.00174 (0.0537, -0.0572) | 0.942 |
| IVH AUC | 0.732 (0.614, 0.851) | 0.806 (0.703, 0.909) | 0.0736 (0.15149, -0.00433) | 0.0605 |
| IVH Sens | 0.44 (0.250, 0.630) | 0.55 (0.348, 0.752) | 0.11 (0.2793, -0.0593) | 0.17 |
| IVH Spec | 0.985 (0.973, 0.996) | 0.971 (0.933, 1.000) | -0.0138 (0.026, -0.0537) | 0.416 |
| Infarct AUC | 0.713 (0.579, 0.848) | 0.823 (0.708, 0.937) | 0.11 (0.1598, 0.0593) | <0.001 |
| Infarct Sens | 0.520 (0.308, 0.732) | 0.776 (0.610, 0.942) | 0.256 (0.5044, 0.0076) | 0.0452 |
| Infarct Spec | 0.850 (0.685, 1.000) | 0.752 (0.619, 0.885) | -0.0976 (0.0328, -0.228) | 0.112 |
| ME AUC | 0.701 (0.596, 0.806) | 0.796 (0.722, 0.870) | 0.0947 (0.1743, 0.0151) | 0.0264 |
| ME Sens | 0.354 (0.131, 0.577) | 0.579 (0.451, 0.708) | 0.225 (0.4343, 0.0157) | 0.0389 |
| ME Spec | 0.978 (0.932, 1.000) | 0.963 (0.931, 0.995) | -0.0157 (0.0268, -0.0582) | 0.393 |
| MLS AUC | 0.891 (0.809, 0.973) | 0.915 (0.790, 1.000) | 0.0239 (0.159, -0.111) | 0.684 |
| MLS Sens | 0.706 (0.472, 0.940) | 0.859 (0.665, 1.000) | 0.153 (0.437, -0.131) | 0.242 |
| MLS Spec | 0.962 (0.874, 1.000) | 0.914 (0.868, 0.961) | -0.0481 (0.0104, -0.1067) | 0.0962 |
| Fracture AUC | 0.769 (0.621, 0.917) | 0.892 (0.815, 0.969) | 0.123 (0.24559, -0.00021) | 0.0503 |
| Fracture Sens | 0.391 (0.0481, 0.7337) | 0.673 (0.5048, 0.8407) | 0.282 (0.6064, -0.0427) | 0.0767 |
| Fracture Spec | 0.983 (0.967, 0.999) | 0.991 (0.980, 1.000) | 0.00781 (0.01945, -0.00382) | 0.156 |

Table S8: Radiographer subgroup

| **Analysis** | **Without_AI** | **With_AI** | **Difference** | **p value** |
| --- | --- | --- | --- | --- |
| Abnormality AUC | 0.886 (0.851, 0.920) | 0.898 (0.862, 0.934) | 0.0123 (0.0505, -0.0259) | 0.5 |
| Abnormality Sens | 0.833 (0.762, 0.904) | 0.905 (0.855, 0.955) | 0.0722 (0.1209, 0.0234) | 0.00674 |
| Abnormality Spec | 0.860 (0.783, 0.938) | 0.809 (0.724, 0.895) | -0.0509 (0.00609, -0.10798) | 0.0775 |
| ICH AUC | 0.832 (0.750, 0.914) | 0.955 (0.928, 0.982) | 0.123 (0.2063, 0.0403) | 0.00724 |
| ICH Sens | 0.858 (0.774, 0.941) | 0.927 (0.879, 0.974) | 0.069 (0.13304, 0.00498) | 0.0368 |
| ICH Spec | 0.965 (0.940, 0.989) | 0.947 (0.911, 0.983) | -0.0177 (0.01, -0.0455) | 0.2 |
| EDH AUC | 0.842 (0.730, 0.954) | 0.876 (0.798, 0.953) | 0.0339 (0.1025, -0.0348) | 0.32 |
| EDH Sens | 0.608 (0.409, 0.807) | 0.700 (0.523, 0.877) | 0.0923 (0.2199, -0.0353) | 0.148 |
| EDH Spec | 0.980 (0.965, 0.996) | 0.975 (0.954, 0.997) | -0.00511 (0.00493, -0.01515) | 0.297 |
| SDH AUC | 0.788 (0.707, 0.869) | 0.855 (0.794, 0.916) | 0.0669 (0.14364, -0.00977) | 0.0829 |
| SDH Sens | 0.521 (0.353, 0.688) | 0.688 (0.561, 0.816) | 0.168 (0.321, 0.0143) | 0.0346 |
| SDH Spec | 0.972 (0.951, 0.992) | 0.959 (0.936, 0.983) | -0.0121 (0.01, -0.0341) | 0.262 |
| SAH AUC | 0.846 (0.767, 0.924) | 0.912 (0.867, 0.957) | 0.0665 (0.1213, 0.0116) | 0.0225 |
| SAH Sens | 0.633 (0.432, 0.834) | 0.826 (0.715, 0.936) | 0.192 (0.3429, 0.0417) | 0.0174 |
| SAH Spec | 0.959 (0.936, 0.981) | 0.919 (0.875, 0.963) | -0.0396 (-0.0123, -0.067) | 0.00626 |
| IPH AUC | 0.852 (0.784, 0.919) | 0.861 (0.803, 0.919) | 0.00914 (0.0594, -0.0411) | 0.709 |
| IPH Sens | 0.737 (0.613, 0.861) | 0.711 (0.583, 0.839) | -0.0257 (0.0697, -0.1211) | 0.581 |
| IPH Spec | 0.940 (0.902, 0.978) | 0.943 (0.905, 0.980) | 0.00261 (0.0436, -0.0383) | 0.893 |
| IVH AUC | 0.825 (0.742, 0.908) | 0.857 (0.782, 0.932) | 0.0322 (0.0832, -0.0188) | 0.214 |
| IVH Sens | 0.615 (0.438, 0.792) | 0.680 (0.523, 0.837) | 0.065 (0.1642, -0.0342) | 0.195 |
| IVH Spec | 0.987 (0.977, 0.997) | 0.985 (0.970, 1.000) | -0.00154 (0.0114, -0.0145) | 0.806 |
| Infarct AUC | 0.797 (0.730, 0.864) | 0.846 (0.787, 0.906) | 0.0495 (0.09039, 0.00865) | 0.0195 |
| Infarct Sens | 0.624 (0.491, 0.757) | 0.732 (0.597, 0.867) | 0.108 (0.1797, 0.0363) | 0.00563 |
| Infarct Spec | 0.904 (0.851, 0.957) | 0.876 (0.812, 0.940) | -0.028 (0.00411, -0.06011) | 0.0827 |
| ME AUC | 0.839 (0.784, 0.895) | 0.844 (0.791, 0.896) | 0.00413 (0.0452, -0.037) | 0.828 |
| ME Sens | 0.706 (0.569, 0.843) | 0.723 (0.589, 0.857) | 0.0167 (0.0829, -0.0496) | 0.583 |
| ME Spec | 0.922 (0.872, 0.971) | 0.912 (0.853, 0.971) | -0.0098 (0.0251, -0.0447) | 0.559 |
| MLS AUC | 0.934 (0.871, 0.997) | 0.945 (0.901, 0.988) | 0.0108 (0.0606, -0.039) | 0.651 |
| MLS Sens | 0.859 (0.722, 0.996) | 0.900 (0.791, 1.000) | 0.0412 (0.1352, -0.0529) | 0.359 |
| MLS Spec | 0.974 (0.955, 0.992) | 0.944 (0.911, 0.978) | -0.0293 (-0.00388, -0.05477) | 0.025 |
| Fracture AUC | 0.858 (0.769, 0.947) | 0.904 (0.846, 0.962) | 0.0459 (0.1244, -0.0327) | 0.225 |
| Fracture Sens | 0.600 (0.392, 0.808) | 0.745 (0.603, 0.888) | 0.145 (0.2606, 0.0303) | 0.0186 |
| Fracture Spec | 0.972 (0.956, 0.988) | 0.981 (0.964, 0.998) | 0.00937 (0.02394, -0.00519) | 0.196 |

Table S9: Senior subgroup

| **Analysis** | **Without_AI** | **With_AI** | **Difference** | **p value** |
| --- | --- | --- | --- | --- |
| Abnormality AUC | 0.896 (0.845, 0.946) | 0.912 (0.876, 0.948) | 0.0165 (0.0561, -0.0232) | 0.385 |
| Abnormality Sens | 0.869 (0.806, 0.932) | 0.905 (0.847, 0.964) | 0.0361 (0.0739, -0.0017) | 0.0597 |
| Abnormality Spec | 0.828 (0.726, 0.931) | 0.794 (0.660, 0.929) | -0.034 (0.0756, -0.1435) | 0.504 |
| ICH AUC | 0.885 (0.831, 0.939) | 0.969 (0.947, 0.991) | 0.0837 (0.142, 0.0254) | 0.00752 |
| ICH Sens | 0.885 (0.817, 0.952) | 0.930 (0.877, 0.982) | 0.0451 (0.08198, 0.00816) | 0.018 |
| ICH Spec | 0.966 (0.940, 0.991) | 0.951 (0.895, 1.000) | -0.0152 (0.0273, -0.0577) | 0.444 |
| EDH AUC | 0.908 (0.842, 0.974) | 0.913 (0.853, 0.973) | 0.005 (0.0249, -0.0149) | 0.586 |
| EDH Sens | 0.777 (0.634, 0.920) | 0.785 (0.631, 0.938) | 0.00769 (0.108, -0.0926) | 0.868 |
| EDH Spec | 0.975 (0.956, 0.994) | 0.980 (0.959, 1.000) | 0.00438 (0.01332, -0.00456) | 0.297 |
| SDH AUC | 0.824 (0.739, 0.908) | 0.905 (0.861, 0.949) | 0.0812 (0.1467, 0.0158) | 0.0183 |
| SDH Sens | 0.536 (0.359, 0.712) | 0.729 (0.615, 0.844) | 0.194 (0.309, 0.0781) | 0.00267 |
| SDH Spec | 0.973 (0.953, 0.994) | 0.960 (0.933, 0.988) | -0.0129 (0.0109, -0.0368) | 0.278 |
| SAH AUC | 0.891 (0.807, 0.976) | 0.951 (0.922, 0.980) | 0.0597 (0.13, -0.0107) | 0.0883 |
| SAH Sens | 0.744 (0.560, 0.927) | 0.908 (0.835, 0.980) | 0.164 (0.3028, 0.0254) | 0.0251 |
| SAH Spec | 0.945 (0.906, 0.984) | 0.913 (0.848, 0.977) | -0.0324 (0.00645, -0.07131) | 0.0955 |
| IPH AUC | 0.885 (0.825, 0.945) | 0.858 (0.795, 0.922) | -0.0265 (0.0246, -0.0776) | 0.303 |
| IPH Sens | 0.783 (0.680, 0.885) | 0.671 (0.540, 0.803) | -0.111 (-0.0227, -0.2002) | 0.0143 |
| IPH Spec | 0.924 (0.882, 0.966) | 0.956 (0.925, 0.986) | 0.0313 (0.06466, -0.00205) | 0.0646 |
| IVH AUC | 0.862 (0.787, 0.937) | 0.828 (0.736, 0.920) | -0.0345 (0.0298, -0.0987) | 0.283 |
| IVH Sens | 0.625 (0.448, 0.802) | 0.610 (0.425, 0.795) | -0.015 (0.0727, -0.1027) | 0.724 |
| IVH Spec | 0.988 (0.972, 1.000) | 0.986 (0.970, 1.000) | -0.00154 (0.00767, -0.01075) | 0.732 |
| Infarct AUC | 0.828 (0.738, 0.918) | 0.860 (0.796, 0.924) | 0.0319 (0.0883, -0.0246) | 0.244 |
| Infarct Sens | 0.668 (0.498, 0.838) | 0.768 (0.642, 0.894) | 0.1 (0.20365, -0.00365) | 0.0572 |
| Infarct Spec | 0.903 (0.837, 0.969) | 0.853 (0.751, 0.954) | -0.0504 (0.00911, -0.10991) | 0.0894 |
| ME AUC | 0.863 (0.809, 0.916) | 0.884 (0.831, 0.937) | 0.0213 (0.0574, -0.0149) | 0.239 |
| ME Sens | 0.738 (0.615, 0.860) | 0.775 (0.662, 0.888) | 0.0375 (0.1074, -0.0324) | 0.267 |
| ME Spec | 0.908 (0.859, 0.957) | 0.906 (0.846, 0.966) | -0.00196 (0.0303, -0.0342) | 0.9 |
| MLS AUC | 0.959 (0.919, 0.999) | 0.962 (0.923, 1.000) | 0.00274 (0.0236, -0.0181) | 0.794 |
| MLS Sens | 0.900 (0.800, 1.000) | 0.941 (0.874, 1.000) | 0.0412 (0.09007, -0.00771) | 0.0893 |
| MLS Spec | 0.968 (0.945, 0.990) | 0.940 (0.902, 0.977) | -0.0278 (0.00189, -0.05753) | 0.0649 |
| Fracture AUC | 0.885 (0.827, 0.944) | 0.918 (0.864, 0.972) | 0.0331 (0.0774, -0.0111) | 0.124 |
| Fracture Sens | 0.664 (0.463, 0.864) | 0.741 (0.585, 0.897) | 0.0773 (0.2078, -0.0532) | 0.222 |
| Fracture Spec | 0.975 (0.961, 0.989) | 0.983 (0.967, 0.998) | 0.00781 (0.02003, -0.00441) | 0.207 |

Table S10: Middle subgroup

| **Analysis** | **Without_AI** | **With_AI** | **Difference** | **p value** |
| --- | --- | --- | --- | --- |
| Abnormality AUC | 0.867 (0.823, 0.912) | 0.887 (0.846, 0.927) | 0.0194 (0.0698, -0.0309) | 0.426 |
| Abnormality Sens | 0.811 (0.750, 0.873) | 0.886 (0.827, 0.945) | 0.0742 (0.124, 0.024) | 0.00561 |
| Abnormality Spec | 0.783 (0.680, 0.886) | 0.734 (0.599, 0.869) | -0.0491 (0.0628, -0.1609) | 0.362 |
| ICH AUC | 0.876 (0.829, 0.923) | 0.947 (0.918, 0.975) | 0.0709 (0.1118, 0.0299) | <0.001 |
| ICH Sens | 0.803 (0.723, 0.883) | 0.903 (0.845, 0.961) | 0.1 (0.1625, 0.0375) | 0.00262 |
| ICH Spec | 0.957 (0.928, 0.986) | 0.919 (0.860, 0.978) | -0.038 (0.00887, -0.08482) | 0.107 |
| EDH AUC | 0.829 (0.720, 0.938) | 0.862 (0.769, 0.955) | 0.0334 (0.1087, -0.0419) | 0.38 |
| EDH Sens | 0.646 (0.470, 0.822) | 0.654 (0.442, 0.866) | 0.00769 (0.149, -0.134) | 0.914 |
| EDH Spec | 0.974 (0.956, 0.993) | 0.979 (0.959, 0.999) | 0.00438 (0.01355, -0.00479) | 0.33 |
| SDH AUC | 0.737 (0.660, 0.814) | 0.871 (0.810, 0.932) | 0.134 (0.211, 0.057) | <0.001 |
| SDH Sens | 0.359 (0.219, 0.499) | 0.694 (0.566, 0.822) | 0.335 (0.44, 0.231) | <0.001 |
| SDH Spec | 0.990 (0.982, 0.998) | 0.957 (0.928, 0.986) | -0.0328 (-0.00758, -0.05794) | 0.011 |
| SAH AUC | 0.787 (0.711, 0.864) | 0.899 (0.859, 0.939) | 0.112 (0.185, 0.0381) | 0.00592 |
| SAH Sens | 0.492 (0.318, 0.667) | 0.800 (0.694, 0.906) | 0.308 (0.455, 0.16) | <0.001 |
| SAH Spec | 0.937 (0.899, 0.974) | 0.878 (0.813, 0.944) | -0.0586 (-0.0191, -0.098) | 0.00442 |
| IPH AUC | 0.829 (0.758, 0.901) | 0.846 (0.784, 0.909) | 0.0169 (0.0772, -0.0434) | 0.577 |
| IPH Sens | 0.703 (0.584, 0.821) | 0.660 (0.523, 0.797) | -0.0429 (0.0794, -0.1651) | 0.479 |
| IPH Spec | 0.906 (0.863, 0.949) | 0.930 (0.889, 0.972) | 0.0243 (0.0744, -0.0257) | 0.319 |
| IVH AUC | 0.782 (0.690, 0.874) | 0.809 (0.710, 0.908) | 0.0269 (0.0922, -0.0385) | 0.414 |
| IVH Sens | 0.480 (0.291, 0.669) | 0.555 (0.367, 0.743) | 0.075 (0.1706, -0.0206) | 0.121 |
| IVH Spec | 0.983 (0.967, 0.999) | 0.979 (0.959, 0.999) | -0.00385 (0.00922, -0.01691) | 0.554 |
| Infarct AUC | 0.774 (0.704, 0.845) | 0.832 (0.766, 0.899) | 0.058 (0.116279, -0.000279) | 0.0511 |
| Infarct Sens | 0.572 (0.425, 0.719) | 0.708 (0.562, 0.854) | 0.136 (0.2459, 0.0261) | 0.0168 |
| Infarct Spec | 0.859 (0.790, 0.928) | 0.795 (0.703, 0.887) | -0.064 (-0.00807, -0.11993) | 0.0274 |
| ME AUC | 0.814 (0.757, 0.870) | 0.853 (0.796, 0.909) | 0.0389 (0.0777, 3.78e-05) | 0.0498 |
| ME Sens | 0.617 (0.500, 0.734) | 0.685 (0.570, 0.801) | 0.0687 (0.14141, -0.00391) | 0.0624 |
| ME Spec | 0.938 (0.910, 0.967) | 0.936 (0.898, 0.975) | -0.00196 (0.0336, -0.0375) | 0.913 |
| MLS AUC | 0.940 (0.887, 0.993) | 0.956 (0.915, 0.997) | 0.0162 (0.0624, -0.03) | 0.476 |
| MLS Sens | 0.859 (0.733, 0.985) | 0.929 (0.826, 1.000) | 0.0706 (0.1686, -0.0274) | 0.154 |
| MLS Spec | 0.968 (0.947, 0.990) | 0.923 (0.883, 0.962) | -0.0459 (-0.0123, -0.0794) | 0.00807 |
| Fracture AUC | 0.811 (0.737, 0.884) | 0.894 (0.829, 0.958) | 0.0829 (0.16, 0.006) | 0.0365 |
| Fracture Sens | 0.432 (0.254, 0.610) | 0.650 (0.491, 0.809) | 0.218 (0.3677, 0.0686) | 0.00596 |
| Fracture Spec | 0.968 (0.951, 0.985) | 0.983 (0.966, 1.000) | 0.0148 (0.03087, -0.00118) | 0.0692 |

Table S11: ED Middle + Senior subgroup
